# Risk of adverse COVID-19 outcomes for people living with HIV: a rapid review and meta-analysis

**DOI:** 10.1101/2020.09.22.20199661

**Authors:** Maya M. Mellor, Anne C. Bast, Nicholas R. Jones, Nia W. Roberts, José M. Ordóñez-Mena, Alastair J.M. Reith, Christopher C. Butler, Philippa C. Matthews, Jienchi Dorward

## Abstract

**Objective:** To assess whether people living with HIV (PLWH) are at increased risk of COVID-19 mortality or adverse outcomes, and whether antiretroviral therapy (ART) influences this risk.

**Design:** Rapid review with meta-analysis and narrative synthesis.

**Methods:** We searched databases including Embase, Medline, medRxiv, and Google Scholar up to 26^th^ August 2020 for studies describing COVID-19 outcomes in PLWH and conducted a meta-analysis of higher quality studies.

**Results:** We identified 1,908 studies and included 19 in the review. In a meta-analysis of five studies, PLWH had a higher risk of COVID-19 mortality (hazard ratio (HR) 1.93, 95% Confidence Interval (CI): 1.59-2.34) compared to people without HIV. Risk of death remained elevated for PLWH in a subgroup analysis of hospitalised cohorts (HR 1.54, 95% CI: 1.05-2.24) and studies of PLWH across all settings (HR 2.08, 95%CI: 1.69-2.56). Eight other studies assessed the association between HIV and COVID-19 outcomes, but provided inconclusive, lower-quality evidence due to potential confounding and selection bias.

There were insufficient data on the effect of CD4+ T cell count and HIV viral load on COVID-19 outcomes. Eleven studies reported COVID-19 outcomes by ART-regimen. In the two largest studies, tenofovir-disoproxil-fumarate (TDF)-based regimens were associated with a lower risk of adverse COVID-19 outcomes, although these analyses are susceptible to confounding by comorbidities.

**Conclusion:** Evidence is emerging that suggests a moderately increased risk of COVID-19 mortality amongst PLWH. Further investigation into the relationship between COVID-19 outcomes and CD4+ T cell count, HIV viral load, ART and the use of TDF is warranted.

## Introduction

By September 2020, over 30 million people worldwide had been diagnosed with severe acute respiratory syndrome coronavirus 2 (SARS-CoV-2)[1]. Although SARS-CoV-2 infection may be asymptomatic or cause only mild symptoms, a proportion of people develop severe coronavirus disease 2019 (COVID-19), leading to hospitalisation, acute respiratory distress syndrome or death. Established risk factors for severe COVID-19 among the general population include older age, chronic kidney disease and obesity [2].

People living with HIV (PLWH), who constitute approximately 0.5% of the global population [3], may have an increased risk of adverse outcomes from COVID-19 as a result of HIV-associated immune dysfunction [4]. There may also be a higher prevalence of comorbidities amongst PLWH that predispose to unfavourable COVID-19 outcomes [5]. Some antiretroviral agents are under consideration as potential treatments for COVID-19 [6], but the influence of antiretroviral therapy (ART) on COVID-19 outcomes is not known. In this rapid review, we aim to evaluate the evidence regarding the risk of adverse COVID-19 outcomes in PLWH, and the extent to which this risk is modified by other factors including ART.

## Methods

We used rapid review methods to identify studies between 1st January and 26th August 2020 that described COVID-19 outcomes in PLWH and compared outcomes with HIV-negative people or the general population, or that compared outcomes by risk factors amongst PLWH. We searched Embase, Medline, medRxiv, LitCovid, Trip, Google and Google Scholar without language restrictions. Search terms are available in Table S1. One author with extensive literature search expertise performed the initial screen to exclude duplicates and studies not related to HIV. For remaining articles, one author performed title and abstract screening, with subsequent full text review by two authors using a standardised data extraction form. Disagreements were resolved by a third author. We included pre-prints in order to capture emerging evidence. Studies with ≤15 participants were excluded as they were unlikely to be powered to detect meaningful associations. We critically appraised the quality of studies using checklists for Case Series and for Cohort Studies from the Joanna Briggs Institute [7].

Cohort studies reporting COVID-19-related death in people with and without HIV that adjusted for age, sex and comorbidities were included in a meta-analysis. Cohort-specific relative risks (RRs) and hazard ratios (HRs) were combined with random effects model to account for variability of the true effect between studies. HRs and RRs numerically approximate each other with shorter follow-up, rarer endpoints, and risks closer to 1 [8]. Subgroup analyses were conducted by study setting and method of confounder adjustment. Meta-analysis was performed in R (version 3.6.0) using the *meta* package [9].

## Results

### Summary of included studies

We identified 1,908 records and included 19 studies in our final qualitative analysis (Figure 1). Seventeen studies [10–26] were peer-reviewed and two were preprints [27,28]. Quality appraisal is included in Tables 1, S2 and S3.

**Table 1:**
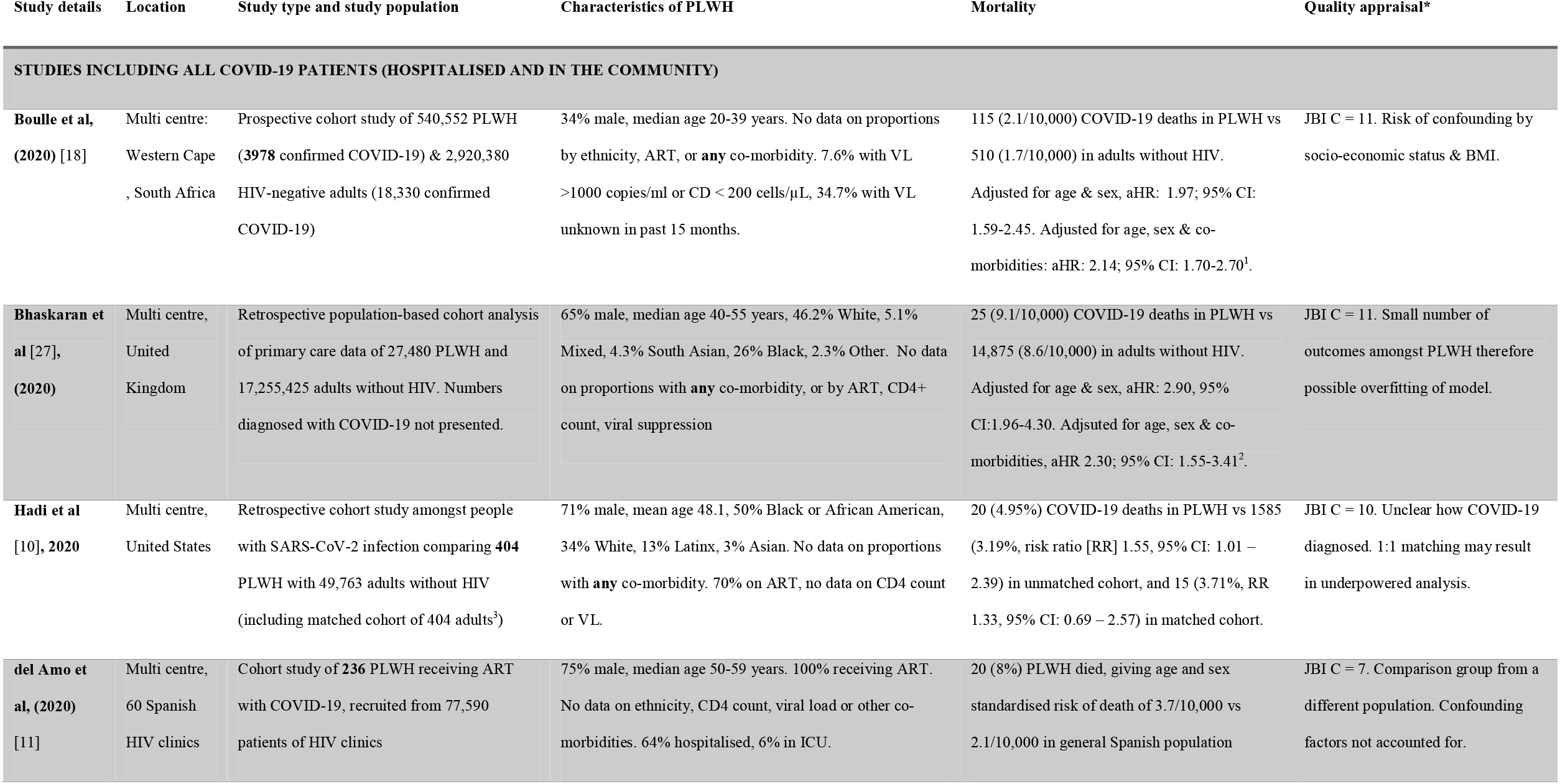
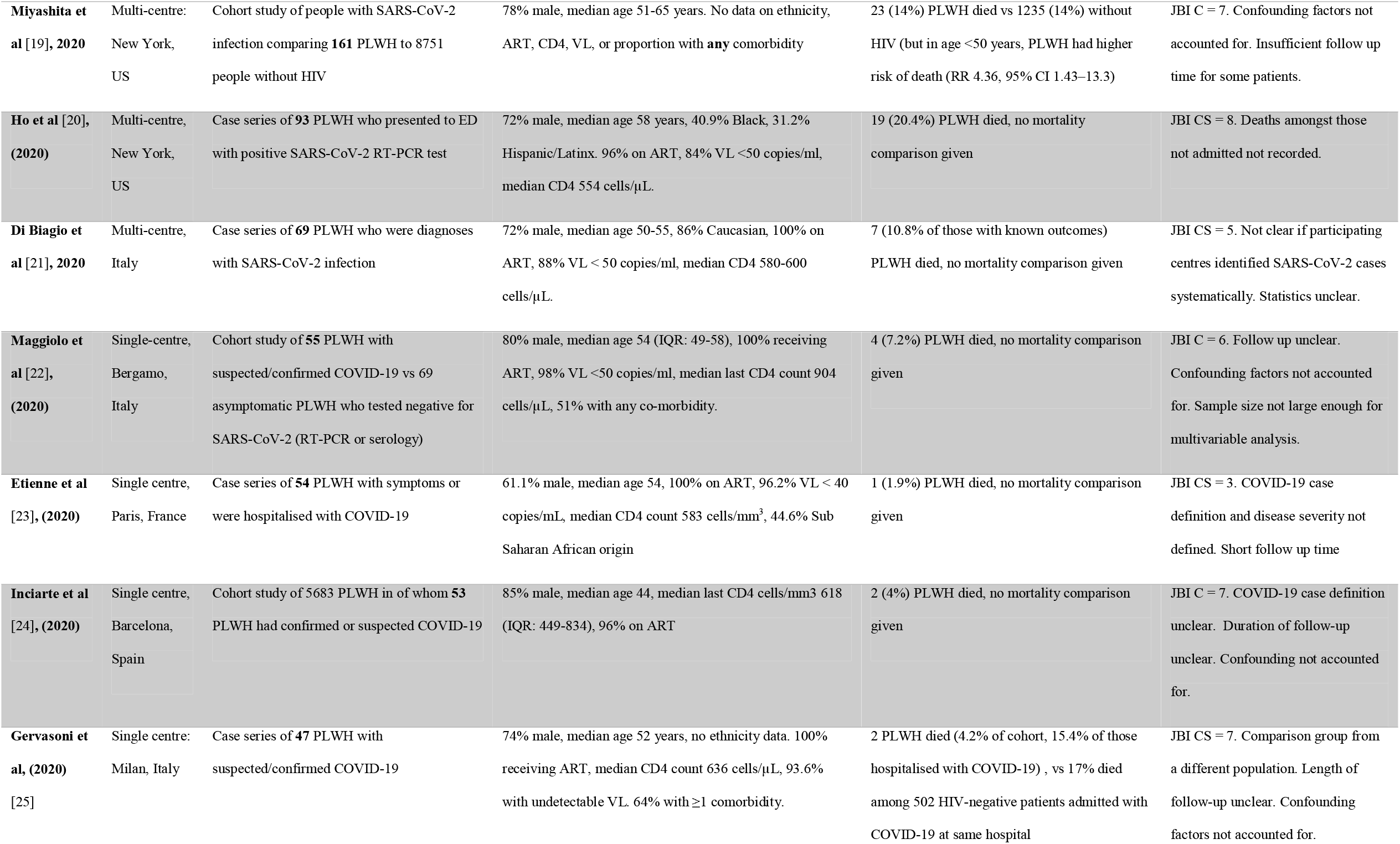
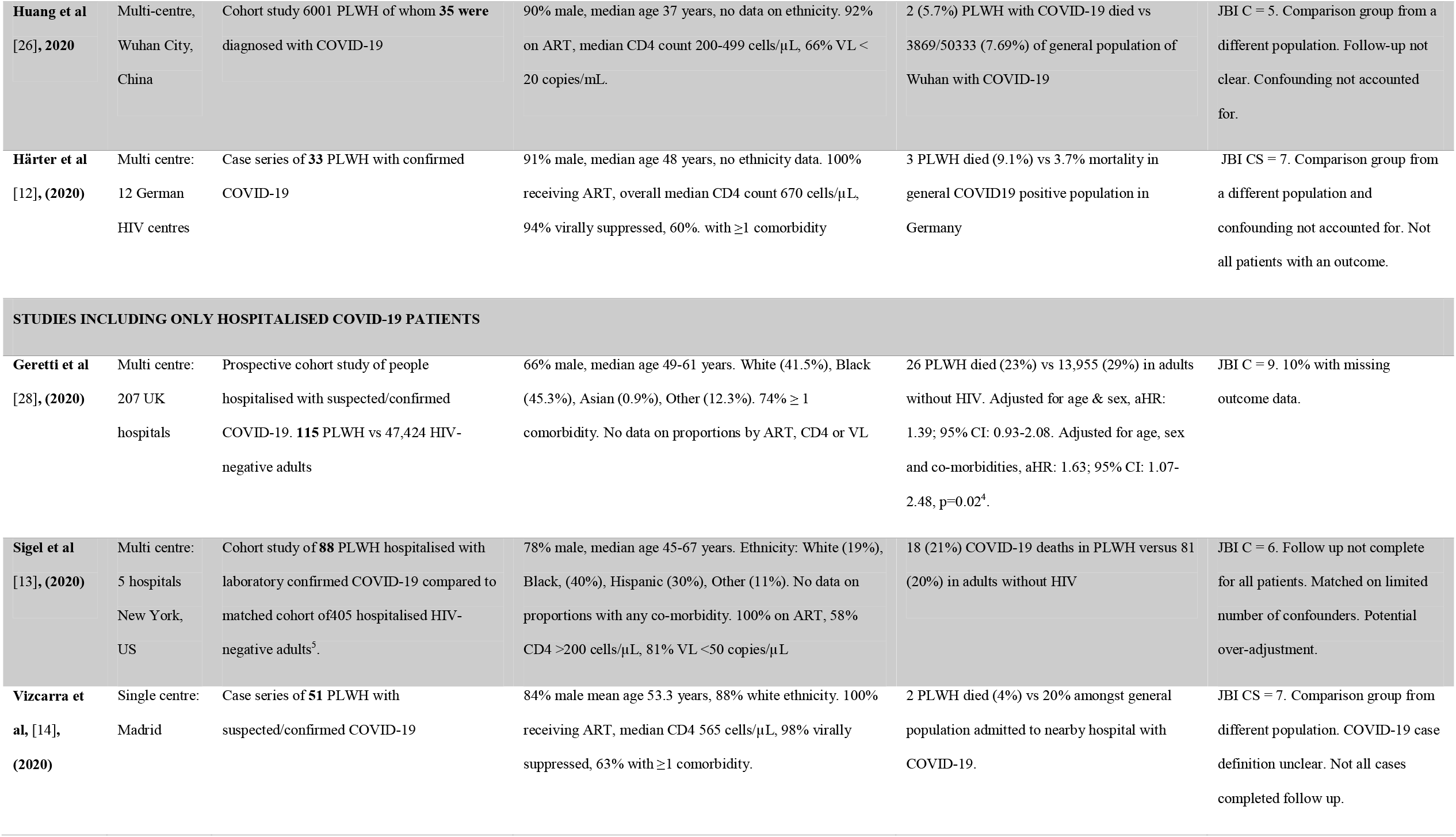
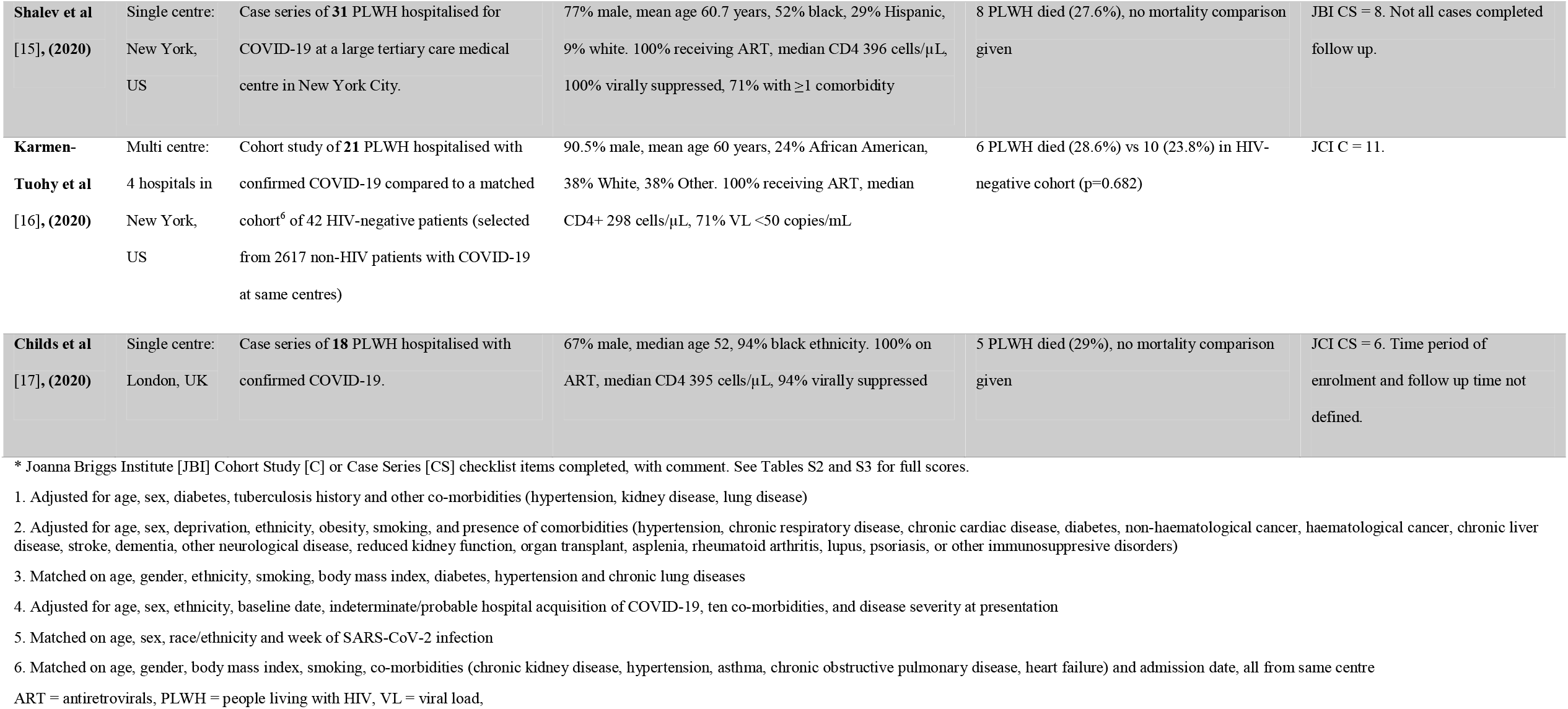
Details of all included studies with summary of mortality findings and quality appraisal

**Figure 1:**
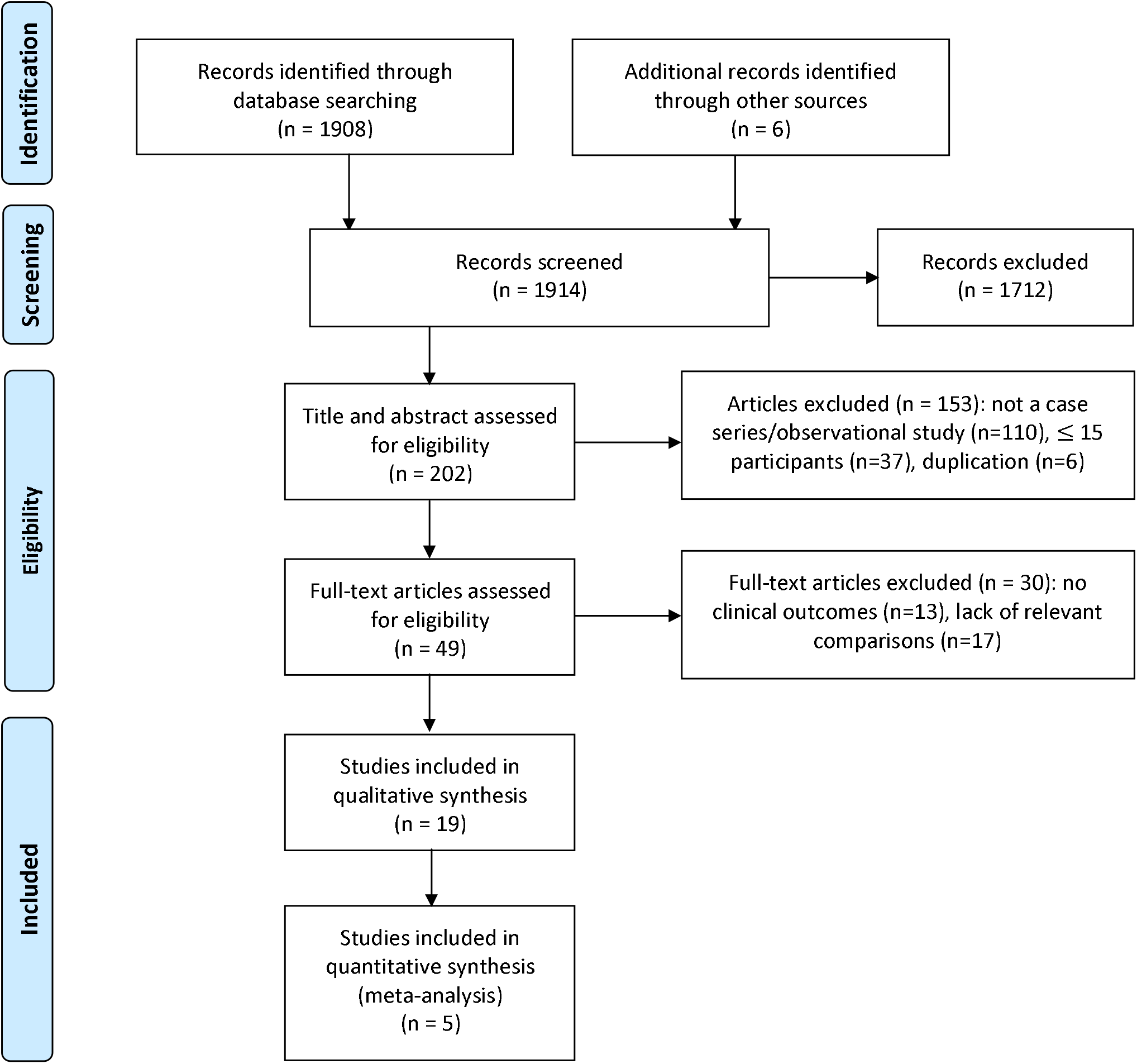
PRISMA Flow diagram to show studies identified and included in a systematic meta-analysis of outcomes of COVID-19 in people living with HIV (PLWH)

**Figure 2.**
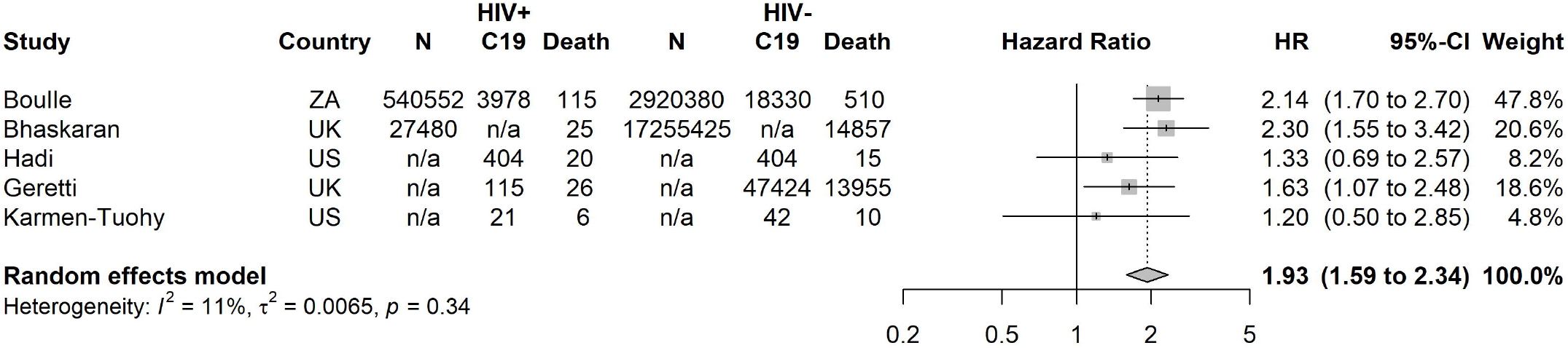
Meta-analysis of the effect of HIV on risk of COVID-19 death. Country; UK = United Kingdom, US = United States of America, ZA = South Africa. HIV+ and HIV-refers to people with and without HIV. C19 refers to those with COVID-19. The denominators (N) refer to the original population where reported: A cohort of people diagnosed with HIV and the wider general population without HIV. Not all studies reported this information. Results are reported as hazard ratios (HR) with 95% confidence intervals (95%CI).

We identified five cohort studies (two prospective, three retrospective) comparing COVID-19 related mortality between PLWH and HIV-negative people, which we pooled in a meta-analysis [10,16,18,27,28]. Four of these reported all-cause mortality among people diagnosed with COVID-19 [10,16,18,28] and one reported mortality due to COVID-19 as recorded on death certificates [27]. Of the remaining fourteen studies, seven made multiple comparisons between PLWH with COVID-19 and HIV-negative cohorts, and/or the general population, and/or PLWH without COVID-19, [11,12,14,15,23,25,26], two studies compared only to a HIV-negative cohort [13,19], two studies compared only cohorts of PLWH with and without COVID-19 [17,22] and three studies compared only the characteristics of PLWH with different COVID-19 disease severity [20,21,24].

There were ten studies that included a total of >1,000 individuals, but amongst these the median number of PLWH with COVID-19 co-infection was only 55 (interquartile range 35-115). Seventeen of the studies were performed in high-income countries and most included a majority of patients on ART with well-controlled HIV (Table 1).

### Quality of evidence and risk of bias assessment

There were common limitations among the included studies. Most were retrospective analyses of routinely-collected clinical data, meaning identification of COVID-19 cases was not systematic and depended on the local approach to screening and diagnosis. This has varied over time and between settings, and may also differ between PLWH and the general population. Only five studies directly compared COVID-19 outcomes amongst PLWH and HIV-negative people in the same cohort, and accounted adequately for potential confounding by co-morbidities associated with adverse COVID-19 outcomes. Other studies used inadequately matched HIV-negative controls, or the general population, which is susceptible to bias as the exposed and control groups were selected differently. Across all studies, the numbers of PLWH and COVID-19 infection were relatively low.

### Adjusted analyses of HIV and risk of death in COVID-19

In a meta-analysis of five cohort studies which accounted for confounding, the risk of death from COVID-19 for PLWH was almost double that of HIV-negative people (HR=1.93, 95% confidence interval [CI] 1.59-2.34) (Figure 1) [10,16,18,27,28]. Three of these studies used large routine databases to identify PLWH across community and hospital settings, in South Africa [18], the United Kingdom (UK)[27] and United States of America (USA)[10], and two studies were limited to hospitalised PLWH and COVID-19 in the UK [28] and USA [16]. In a subgroup analysis there was no significant difference between study settings (p=0.17), although a weaker HR was seen in hospitalised patients (Figure S1). Among the three studies which used multivariable adjustment to account for confounding [18,27,28], the *crude* risk of COVID-19 death was similar between people with and without HIV, but after adjustment for age, the adjusted risk amongst PLWH was higher. Subsequent adjustment for comorbidities did not drastically alter HRs (Table 1). In sub-group analysis by method of accounting for confounders, a weaker HR was seen in the smaller two studies which used propensity score matching (Figure S2) [10,16].

### Adjusted analyses of risk of hospitalisation and morbidity in PLWH

Three of the five cohort studies conducted analyses of the association between HIV status and the risk of other COVID-19 outcomes [10,16,28]. Amongst 47,539 individuals hospitalised with COVID-19 in the UK, the odds of critical care admission was higher among the 115 PLWH (odds ratio (OR) 2.60, 95 % CI 1.74-3.87), but this was attenuated after adjustment for demographics and co-morbidities (aOR 1.13, 95% CI 0.72-1.75) [28]. In the analysis of 50,167 people with COVID-19 in the USA, 404 PLWH (n=404) were at increased risk of hospitalisation compared to 1:1 propensity score matched HIV-negative controls (RR 1.70, 95% CI: 1.21-2.38) [10]. Lastly, among 2,638 people hospitalised with COVID-19 in the USA, there were 6 (28.6%) intensive care unit admissions among 21 PLWH, compared to 7 (16.7%) amongst a propensity score matched cohort of 42 HIV negative people (p = 0.271)[16].

### Unadjusted analyses of COVID-19 related outcomes in PLWH

Eight studies provided lower-quality evidence regarding COVID-19 outcomes in PLWH as they did not compare HIV-positive and negative people in the same cohort, or did not adequately account for confounders. The largest of these was a Spanish multi-centre study of 77,590 PLWH, of whom 236 were diagnosed with COVID-19 and 20 died. In keeping with our meta-analysis result, age- and sex-standardised mortality from COVID-19 were found to be higher in PLWH (3.7 per 10,000) compared to the general population (2.1 per 10,000) [11]. The other seven studies [12–15,19,25,26] were limited due to being at single sites, having small sample sizes (median 64, range n=31-161 PLWH diagnosed with COVID-19) and not accounting for potential confounding. These studies report conflicting results with one suggesting a higher rate of hospitalisation and mortality among PLWH compared to the general COVID-19-positive population [12], two studies suggesting lower COVID-19 mortality in PLWH [14,25] and four studies reporting no significant difference in the risk of adverse outcomes from COVID-19 between PLWH and HIV-negative cohorts [13,19] or the general population [15,26].

### Risk of death and hospitalisation in relation to CD4+ T cell count and HIV viral load

Several of the large cohort studies did not include data on CD4+ T cell count or HIV viral loads [10,27,28]. In the South African study, lower CD4+ T cell counts (measured during the COVID-19 episode) were associated with mortality, but this could be a result of, rather than causing, severe disease (Table 2). There was no difference in outcomes by HIV viral load, although viral load data was incomplete and numbers with unsuppressed viral loads were small [18]. A London HIV clinic found that 18 PLWH who were hospitalised with COVID-19 had a lower median CD4+ T cell count (395 vs 573 cells/µL, p=0.03) compared to their 2,699 PLWH outpatients (Table 2) [17]. A further nine studies (median n= 54 PLWH and COVID-19, range n=35-93) found no significant association between CD4+ T cell count or HIV viral load and COVID-19 outcomes [13,14,16,20–24,26].

**Table 2:** Summary of studies reporting outcomes by ART regimen and other risk factors for adverse COVID-19 outcomes amongst people living with HIV

| STUDY | INFLUENCE OF ART REGIMENS | OTHER RISK FACTORS AMONG PLWH |
| --- | --- | --- |
| **Boulle et al, (2020)**<br>[18]n=3978 | Lower mortality in patients on TDF vs abacavir/zidovudine (aHR: 0.42; 95% CI: 0.22;0.78) | 601 patients had CD4 count measured during episode of C-19. Higher mortality associated with CD4 counts <200 cells/μL (n=70) vs ≥350 cells/μL (aHR 1.97; 95% CI 1.14-3.40). Direction of causality unclear.<br>No difference in hazard of C-19 death by HIV VL (aHR vs HIV-negative: 2.61 (95% CI: 1.98-3.43) for VL < 1000 copies/mL; 3.35 (95% CI: 1.38-6.12) for VL ≥ 1000 copies/mL or CD4 <200 cells/μL). |
| **Bhaskaran et al [27], (2020)**<br>**Number of PLWH & COVID-19 not presented** | N/A | PLWH of Black ethnicity had higher risk of COVID-19 mortality (aHR 3.80, p for interaction 0.045). No data on outcomes by CD4 or VL. |
| **del Amo et al, (2020) [11]**<br>**n=236** | Lowest risk for C-19 diagnosis (16.9% [95% CI: 10.5 - 25.9] & hospitalisation (10.5 [95% CI: 5.6 - 17.9] ) in PLWH receiving TDF/FTC compared to other ART regimens (e.g. ABC/3TC 28.3% [95% CI: 21.5 - 36.7] and 23.4% [95% CI: 17.2 - 31.1] respectively). | Higher crude risk of C-19 death amongst older PLWH (70-79 years = 26.6/10,000, 95% [CI 10.7–54.9] vs 50-59 years 2.2/10,000 [95% CI 10.7-54.9]). No difference by sex. |
| **Ho et al [20], (2020)**<br>**n=93** | No significant difference in TDF use between PLWH with C-19 who survived and died (73.6% vs 55.5%, p=0.15). | No significant differences in obesity, CD4 counts or HIV VL between PLWH with C-19 who survived and died |
| **Di Biagio et al [21], 2020**<br>**n=69** | No stat. sig. association between risk of hospitalisation and ART regimens | Hospitalised PLWH were slightly older (p=0.047). No association between most recent VL or CD4 count and hospitalisation. |
| **Maggiolo et al [22], (2020)**<br>**n=55** | No difference in TDF use amongst PLWH with C-19 (60%) vs without C-19 (60.8%) | 4 PLWH with C-19 who died had lower last CD4 count (median 514 cells/μL) than the 51 PLWH who survived (median 913 cells/μL). |
| **Etienne et al [23], (2020)**<br>**n=54** | No stat. sig. difference between ART regimen and C-19 severity | No stat. sig. association between CD4 counts or VL < 40 copies/mL and C-19 severity |
| **Inciarte et al [24], (2020)**<br>**n=53** | No associations between ART regimen and C-19 severity | No association between latest CD4 count and C-19 severity |
| **Huang et al [26], 2020**<br>**n=35** | N/A | Older age and ART discontinuation associated with C-19 infection. No association between latest CD4 count or VL and C-19 infection. |
| **Geretti et al [28], (2020)**<br>**n=115** | N/A | Age, obesity and diabetes were associated with C-19 death amongst PLWH. |
| **Sigel et al [13], (2020)**<br>**n=88** | PLWH who survived were more likely to have been treated with NRTIs than those PLWH who died (99% v 89%, p=0.04) in univariate analysis. No difference in outcomes for other classes of ART. | No association between comorbidities, latest CD4 count or VL and C-19 death. |
| **Vizcarra et al, [14], (2020)**<br>**n=51** | Increased TAF use in PLWH with C-19 (37/51, 73%), vs PLWH without COVID-19 (38%, p=0.0036) | PLWH with C-19 were significantly more likely to have comorbidities (63% vs 38%, P=0.00059), and had higher median BMI (25.5 kg/m<sup>2</sup> vs 23.7 kg/m<sup>2</sup>, P=0.021) compared to 1288 PLWH without C-19. No association between CD4+ T cell count and SARS-CoV-2 infection or adverse C-19 outcomes. |
| **Shalev et al [15], (2020)**<br>**n=31** | 7/8 (88%) PLWH who died from C-19 used TAF/TDF vs 10/23 (43%) of those who survived | N/A |
| **Karmen-Tuohy et al [16], (2020)**<br>**n=21** | N/A | No association between most recent CD4 count and mortality (OR 0.996, 95% CI: 0.992 to 1.11) |
| **Childs et al [17], (2020)**<br>**n=18** | More common use of protease inhibitor-containing ART regimens among PLWH with C-19 (OR, 2.43, 95% CI, .94–6.29) | PLWH hospitalised with C-19 were more likely to be of black ethnicity (OR: 12.22, 95% CI: 1.62-92.00), and had lower median CD4+ cell counts (395 vs 573, P=0.03) |

3TC = lamivudine, ABC = abacavir, FTC = emtricitabine, ART = antiretroviral therapy, BMI = body mass index, C-19 = COVID-19, OR = odds ratio, PLWH = people living with HIV, stat. sig. = statistically significant, TAF = tenofovir alafenamide, TDF = tenofovir disoproxil fumarate, VL = viral load

### Impact of ART regimen on COVID-19 outcomes

No studies compared COVID-19 outcomes between PLWH receiving and not receiving ART. We identified eleven studies assessing the relationship between specific ART regimens and COVID-19 outcomes in PLWH. In South Africa, COVID-19 related mortality was lower in patients on TDF-based regimens versus abacavir/zidovudine-based regimens, which are used for patients with co-morbidities or requiring second line treatment (aHR: 0.42; 95% CI: 0.22-0.78) [18]. While this analysis was adjusted for certain co-morbidities, the observed association may be confounded due to patients receiving TDF having less complex healthcare needs. In the Spanish multi-centre study, PLWH receiving TDF and emtricitabine (FTC) had the lowest risk for COVID-19 diagnosis (16.9 per 10,000) and hospitalisation (10.5 per 10,000) compared to all other ART regimens investigated, but without adjusting for co-morbidities [11]. A US hospital study found that PLWH and COVID-19 who survived were more likely to have been treated with NRTIs that those PLWH who died (99% vs 89%, p=0.04) in univariate analysis [13]. Seven smaller studies (n=18-93 PLWH with COVID-19) reported no significant association between ART-regimen and COVID-19 severity amongst PLWH [14,15,17,20–24].

### Other factors influencing COVID-19 outcomes among PLWH

Thirteen studies assessed the influence of comorbidities and demographics on the outcomes of COVID-19 amongst PLWH [13,14,16–18,20–24,26–28]. In the UK-based cohort study of hospitalised patients, among 115 PLWH with COVID-19, the 26 who died were more likely to have obesity and diabetes [28]. Bhaskaran et al and Childs et al report evidence of a higher risk of COVID-19 death and hospitalisation respectively amongst PLWH of black ethnicity (Bhaskaran et al mortality aHR=3.80; 95% CI: 2.15-6.74, p for interaction=0.045; Childs et al hospitalisation crude OR: 12.22, 95% CI: 1.62-92.00) [17,27]. Other smaller analyses suggested that amongst PLWH, factors such as older age [23], metabolic disorders [23], obesity [14], African ethnicity [23] and organ transplantation [13] were associated with COVID-19 infection or severity.

## Discussion

### Summary

Emerging evidence suggests an increased risk of COVID-19 related death in PLWH. There was insufficient data to determine whether HIV viral load, CD4+ T cell counts or ART use are associated with COVID-19 related death. We found some evidence that among PLWH receiving ART, TDF use may be associated with lower frequency of SARS-CoV-2 infection and milder courses of COVID-19, although this was not consistent between studies and was susceptible to confounding. Risk factors for severe COVID-19 among PLWH include older age, obesity and black ethnicity, and appear similar to the general population.

### Risk of COVID-19 related mortality among PLWH

In our review, the two population-based studies from South Africa and the UK both suggested almost double the risk of COVID-19 related death amongst PLWH, despite having very different demographic profiles [18,27]. In contrast, studies restricted to cohorts of PLWH diagnosed with COVID-19 [10], and hospitalised patients with COVID-19 [16,28] found a weaker or null effect. These studies are more at risk of selection bias, as PLWH with milder symptoms may be more likely to test for SARS-CoV-2 or be hospitalised by clinicians (due to a higher perceived risk), compared to people without HIV who may only be tested or hospitalised once more severely unwell. This would lead to the cohort of PLWH being less unwell at baseline compared to the HIV negative cohort, leading to underestimation of any association between HIV status and COVID-19 related mortality. Furthermore, studies restricted to hospitalised patients cannot account for the effect of HIV (or any other potential risk factor) on SARS-CoV-2 infection and COVID-19 severity which result in hospitalisation, and therefore may underestimate the effect of risk factors on COVID-19 death, compared to studies in the general population [29].

### Influence of ART

We found no evidence to determine whether ART reduces COVID-19 severity through immune reconstitution, as most studies included PLWH on ART. Regarding specific antiretrovirals, the potential therapeutic value of TDF for COVID-19 is supported by results from molecular docking studies [30]. However, TDF is relatively contra-indicated in renal impairment [31], meaning patients receiving TDF-based ART are likely to have less comorbidities, which may explain the observed better COVID-19 outcomes. Randomised trials of TDF prophylaxis for SARS-CoV-2 are underway [32].

### Comparisons with existing literature

PLWH are known to be at higher risk of respiratory bacterial infections, but the evidence regarding acute viral infections is less clear [33]. A review from the H1N1 influenza pandemic in 2009/2010 found some evidence of a higher risk of adverse H1N1 outcomes amongst PLWH who were severely immunocompromised [34]. However, the quality of the evidence was weak with a lack of rigorously-designed prospective cohort studies, reflecting the challenges of in-pandemic research [34].

As of August 26^th^ 2020, we identified seven systematic reviews on COVID-19 in PLWH [35–41]. All these reviews lacked the more robust evidence from recent large cohort studies [18,27,28]. Moreover, one review included articles assessing non-HIV related immunodeficiency [38] and four did not address the influence of ART [35,39–41].

### Limitations

Our meta-analysis of five studies is potentially limited by the small numbers of PLWH with COVID-19 who died. This presented challenges when accounting for confounding; studies that used multivariable analyses to adjust for confounding were susceptible to over-fitting of models and potential over-adjustment by factors which could be on the causal pathway between HIV and death (e.g. malignancy or tuberculosis). Studies that used matching were potentially under-powered, which may explain why they tended to report no independent association between HIV and COVID-19 death. In our narrative synthesis, the majority of the studies were small case series or cohort studies that did not adequately account for confounders such as age. Most were performed in high-income countries, and the majority of participants had well-controlled HIV on ART. This may limit the applicability to populations of PLWH in other settings. Only 68% of adults and 53% of children living with HIV globally are receiving ART [42], highlighting a crucial need to examine the risk of COVID-19 complications in these populations.

## Conclusion

We present evidence which suggests a moderately increased risk of COVID-19 death among PLWH. TDF-based ART may be associated with lower risk of COVID-19 diagnosis and adverse outcomes, although this finding is susceptible to confounding. Measures to mitigate COVID-19 risk among PLWH should be included in the global pandemic response, while further research into the role of ART, immunosuppression and viral suppression is needed to quantify and address risks for PLWH in diverse settings.

## Data Availability

No data available

## Acknowledgements

MM, NJ and JD conceived the study. NR performed the literature search. MM and AB contributed equally to the screening and qualitative analysis, with support from NJ and JD. JOM performed the meta-analysis. MM and AB wrote the first draft of the manuscript. MM, AB, NR, JOM, AR, CB, PM, NJ and JD critically reviewed and edited the manuscript and consented to publication.

